# Determinants of Oral Chemotherapy Adherence: A Social Cognitive Perspective

**DOI:** 10.1101/2025.10.02.25337011

**Authors:** Solomon Ayehu, Michael Novatnack, Bethel Belayneh, Dominic Tecza, Leah L. Zullig, William A. Wood, Marcella H. Boynton, Emily Mackler, Ashley Leak Bryant, Stephanie B. Wheeler, Jennifer Elston Lafata, Benyam Muluneh

## Abstract

**PURPOSE:** Oral anticancer agents (OAAs) have transformed cancer treatment by allowing home-based therapy, but adherence remains suboptimal in real-world practice. Guided by Social Cognitive Theory (SCT), this study examined behavioral, cognitive, and socioenvironmental determinants of OAA adherence.

**PATIENTS and METHODS:** We conducted 36 semi-structured interviews with adult cancer patients receiving OAA monotherapy at an academic medical center, a large urban facility, and a rural facility. Interviews were analyzed using a critical realist approach to thematic analysis, deductively guided by SCT. Themes were organized according to the World Health Organization’s (WHO) dimensions of adherence.

**RESULTS:** Five interrelated themes shaped adherence across WHO dimensions: “[I feel] very confident…I’m staying positive about it”: Knowledge, Skills, and Self-Efficacy; “They’ll stop me from speaking”: Communication and Trust with Providers; “We’re at Their Mercy”: Logistical Factors in Prescription Refills and Shipment; “Who can afford that?”: Distress Related to High Costs and Insurance Coverage Challenges; and “My wife…knows sometimes I forget”: Family and Social Support in OAAs Adherence.

These findings reflected SCT constructs including self-efficacy, behavioral capability, reinforcement, observational learning, and reciprocal determinism, highlighting the dynamic interactions among personal, social, and health system factors in adherence behavior.

**CONCLUSION:** OAA adherence is shaped by multilevel determinants that extend beyond individual barriers to include provider communication, system logistics, financial burden, and social support. Interventions to improve adherence should be theory-informed, multidisciplinary, and patient-centered, addressing behavioral, relational, and structural influences simultaneously.

## Introduction

Oral anticancer agents (OAAs) have transformed cancer treatment by offering the convenience of home-based therapy and reducing the need for hospital visits compared to IV chemotherapy. Adherence to these long-term therapies is essential to achieve optimal clinical outcomes, improve survival, and enhance quality of life, often requiring adherence rates exceeding 90% [1–2]. Despite the advantages of at-home administration, adherence to OAAs in routine practice is often significantly lower than anticipated [3–5]. The shift to OAAs places significant responsibility on individuals to consistently follow complex regimens, monitor side effects, and sustain motivation throughout extended treatment durations. Decades of research on medication adherence consistently find that no matter how life sustaining an oral medication may be, a large proportion of patients find it difficult to maintain high levels of medication adherence, especially with more complex or multi-dose medication regimens [6–7].

Although OAAs offer significant advantages in autonomy and flexibility, they also introduce unique adherence challenges due to reduced face-to-face clinical oversight. In addition to regimen complexity, challenges such as side effects, financial burden, forgetfulness, and potentially limited understanding of treatment importance are all substantial barriers to adherence [3, 8–10]. Prior studies of OAA adherence have mainly focused on proximal barriers (e.g., side effects, forgetfulness) without addressing underlying psychological factors impacting adherence. Few studies on OAA adherence have leveraged psychosocial or behavioral theories to examine drivers of OAA adherence. Lack of theory-based psychosocial understanding of OAA adherence and its barriers limits the ability to design maximally effective adherence interventions.

To address this gap in the research literature, we conducted a qualitative study guided by Bandura’s Social Cognitive Theory (SCT) to explore barriers to OAA adherence and the behavioral, cognitive, and socioenvironmental factors influencing OAA medication taking behavior in real-world settings [11]. SCT is particularly suited for understanding complex health behaviors that require sustained effort, self-regulation, and support systems.

## Methods

### Study Design

We used semi-structured in-depth interviews (IDIs) to gather data from participants identified through the electronic medical records (EMR) system. We followed a deductive approach to reflexive thematic analysis (TA) using Braun and Clarke’s (2006) six-phase framework to TA [12]. The interview guide and interpretation of adherence to OAA findings utilized primary constructs within the SCT. These constructs include self-efficacy, behavioral capability, reinforcement, observational learning, and reciprocal determinism – the dynamic interaction of the person, behavior, and environment in shaping health outcomes [13]. The research procedures conformed to the Declaration of Helsinki and were approved by the UNC Institutional Review Board (IRB 21-0744).

### Study Settings and Participant Eligibility

Our methods have been previously published [14]. Eligible participants were adult patients aged 21 years and older, diagnosed with cancer, taking OAA monotherapy for at least 6 months, and treated at a UNC Health affiliated facility between August 2023 and May 2024 dates. For participants who were not able to interview independently, caregivers were allowed to aid in providing answers. Participants were purposively sampled to ensure proportional gender representation and to meet a 30% quota for minoritized groups, including Black, Hispanic, Asian, and Indigenous populations. Additionally, the sample was reflective of patients on OAA therapy receiving care across the health system’s diverse settings (one academic medical center, one large urban facility, and a rural facility.

### Recruitment and Data Collection

Eligible patients were contacted by phone and invited to participate in a research interview. Once identified and consented, the participant’s contact information was shared with the UNC Connected Health for Applications & Interventions (CHAI) Core Qualitative Research Team. A trained member of CHAI Core team contacted eligible participants and conducted the interview. CHAI Core team members conducting interviews for this study had a background in public health.

The semi-structured interview guide was informed by the SCT and developed collaboratively with the study team, to elicit insights and descriptive details from the participants’ perspectives (see Appendix). Participants received a $50 gift card upon completing the interview. The guide addressed personal, environmental, social, medication factors, prescription filling, outcome expectations, and motivations. The research team pilot tested the interview guide and refined order and phrasing as appropriate. Interviews were audio-recorded and transcribed verbatim by a professional service.

### Data Analysis

We employed a critical realist approach to deductive thematic analysis, guided by the SCT framework. Our analysis evolved from an initially semantic to a more latent conceptualization [15]. Members of the CHAI qualitative research team reviewed each transcript for accuracy and consistency while taking notes to familiarize themselves with the data. A codebook was developed based on key constructs of the SCT, interview questions, and notes taken during data collection. De-identified transcripts were imported into Dedoose (SocioCultural Research Consultants, LLC. Dedoose. https://www.dedoose.com/. Accessed Spring 2024) for data management. The initial codebook was pilot tested in two rounds by independently coding four transcripts, which led to fine-tuning concept definitions and revising decision rules. This process continued until replicability occurred across coders. We followed standard consensus coding guidelines, where any emerging theme or discrepancy was captured and reconciled through discussion and consensus.

We generated initial themes that reflected provisional and broad categorical topics encompassing individual, social, medication, and environmental factors and participants’ final recommendations. In reviewing our themes, we decided to cluster our codes according to the WHO’s five dimensions of adherence: patient-related, health system-related, therapy-related, condition-related, and socioeconomic factors [16]. This structured approach, based on WHO’s taxonomy, facilitated the identification of dominant and deeper emerging themes. We maintained reflexivity throughout this process by engaging in regular discussions within the multidisciplinary study team that included clinical pharmacy fellows and those with expertise in public health, medicine, and health policy, and by keeping reflexive journaling.

Our findings are presented through illustrative quotes, integrating descriptive accounts and interpretative analysis grounded in SCT. The final manuscript adheres to the Standards for Reporting Qualitative Research (SRQR) to ensure methodological transparency and rigor.

## Results

### Participant Characteristics

We conducted 36 individual telephone interviews between December 2023 and April 2024. Participants’ median age was 62 and a half years, with 22 females and 14 males. 12 participants received care at academic medical center, 12 at the large urban community facility, and 12 at the small rural facility. We use “urban” and “rural” as defined by the Office of Management and Budget as adhered to by the U.S. Department of Health and Human Services [17]. One interview was conducted with a caregiver (the participant had dementia), and another interview had two participants (the participant and their adult child). On average, interviews lasted 35 minutes.

### Qualitative findings

Our coding originally produced seventeen broad and semantic codes. We further delineated and classified over a hundred latent subcodes through an iterative coding process. After thoroughly reviewing the codes, coded data, and full dataset, we prioritized latent ideas underpinning key domains of SCT constructs—personal cognitive, behavioral, and socioenvironmental factors. We structured our analysis around five main themes and mapped the themes across three of the relevant WHO’s five dimensions of adherence, which were patient-related, social/economic, and health care team/system-related domains [16].

## Patient-related factors

### Theme 1: “[I feel] very confident…I’m staying positive about it”: Knowledge, Skills, and Self-Efficacy

In discussing personal influences, participants noted barriers and facilitators that they have encountered and highlighted recommendations for how providers could further support treatment adherence.

Patients’ confidence levels in managing OAA-related side effects emerged as a salient factor in their treatment experience. Reported confidence levels varied. Some participants indicated that uncertainties around diagnosis and treatment plans were a significant source of self-doubt throughout the process of taking or managing their medication. One expressed that they “don’t feel very confident” (P12) because “no doctors want to listen to all of the symptoms I’m having” (P12) as side effects. By contrast, another said they are “pretty confident [about managing side effects]” (P11) and are open to asking their providers for assistance because providers are “pretty responsive” (P11). Several participants said they felt “very confident” (P30) in managing communications with the clinic about side effects. One participant expressed this idea in highly self-assured terms, saying, “I haven’t had anything yet that has come across that I haven’t been able to handle. I feel like the medication is really doing its job” (P2). Such variations in confidence around side-effect management likely influence how consistently patients adhere to their prescribed OAA regimens.

Participants’ confidence in managing their OAA was also reflected in their strategies for dealing with routine disruption. For instance, when spending time out of the house or when traveling, one participant described taking the OAA “medication with me always” (P9). Some have also built strategies for stretching medications between shipments by saving extra pills in case of prescription refill delays and other disruptions. One participant described “[keeping] a three- or four-day buffer” for such moments (P16).

Participants also reported knowledge as a key factor for adherence to OAAs. Their understandings of their diagnoses, treatment plans, medication benefits, and regimens all appeared to affect adherence coupled with mixed levels of knowledge. Participants had different understandings of the consequences of skipping OAA doses. Some expressed awareness of the risks of missing doses. One said, “[Patients] may see [fewer] side effects of the drug, but … see more side effects of the disease” (P26). Another indicated concern that non-adherence would “prolong the [wait for] remission” and that they “would never skip it” (P12). One patient emphasized “the importance of [OAA treatment],” acknowledging that “there are side effects, but there are [also] tremendous benefits to being able to have this medication available to us. To be able to take it to make us better was the main reason (P9)” for maintaining adherence to OAAs. By contrast, other participants expressed the belief that missing doses sporadically was not harmful and that only “the leveraged [effect] of doing that multiple times over an extended period …” (P5) was consequential. Finally, some expressed uncertainty, saying they “don’t know” the risks of missing doses or stopping treatment early (P3). One said that they “[did not] understand the difference between why some people have three years [of treatment] and some people have five” (P3). Personal efforts to mitigate forgetfulness included using “a reminder” (P14), “calendar on the phone” (P35), or “paper calendar” (P25) and taking medication as “part of [one’s routine]” (P1).

## Health care team and system-related factors

### Theme 2: “They’ll stop me from speaking”: Communication and Trust with Providers

Patients who struggled with adherence often indicated that their healthcare providers did not ensure their understanding. One participant stated:

> “I would say give more information and research. I’m not sayin’ that you guys don’t. I’m just speakin’ from my experience. To be switched to a medication and say these are common side effects is kinda unfair because you don’t know what side effects the person that’s gonna actually take the medication is gonna have. I think that explainin’ the medication more or at least bein’ able to provide a booklet, a pamphlet, or somethin’ about the medication is more helpful.” (P12).

Indeed, participants described a care delivery system in which providers do not actively mitigate knowledge gaps. Often, these gaps resulted in a lack of understanding for participants who resume a medication regimen after a break, with one patient saying this process “can be very confusing” (P31) and another expressing that they were “uncertain” (P14) about whether to take their medication with food and how to time doses. Another area of confusion was how to adapt their medication regimen to accommodate undergoing reconstructive surgery. Commenting on the level of awareness needed around delays in reconstructive surgery associated with chemotherapy, one participant said: “Some cancer patients are going to need multiple surgeries, so how does this drug affect you, and what would you need to do?’ seems like a very logical conversation for a doctor to have with you…The oncologist, pharmacist, and plastic surgeon all had an excellent opportunity to say something so that I wouldn’t have been [overwhelmed with the news that] I’m [going to] wait for two years to have reconstruction” (P13).

Patients noted that their relationships with providers may significantly impact adherence to OAAs. Most participants identified effective communication with providers as a valuable determinant of adherence. Communication about side effects was significant. One participant described not feeling “very confident” with the level of support they were receiving for side effects, noting: “When something happens, and I’m faced with something, [providers] be like, ‘Well, this is not a side effect, this is not a common side effect,’ [which] makes me feel like … it’s coming from something else. Then I kind of feel like the issues are never addressed.” (P12). Another participant asked, “At what point are [providers] going to actually take me seriously and figure out what’s going on?” (P3). Another described being “passed around by so many specialists because no doctor wants to listen to all of the symptoms I’m having” (P3). Similarly, one participant asserted:

> “… As soon as one of the symptoms I’m telling them [about] is out of their realm [of expertise], they’ll stop me from speaking and say that they’re going to have to send me to someone else. I just wish that someone would listen to me fully and then maybe have a suggestion [for] someone [who] could look at me as a holistic part. I’m so sick of—it’s been a year plus of me just going—bouncing around from specialist to specialist with absolutely no help and not getting out of any pain. I think that’s just driving me insane” (P3).

By contrast, some participants appreciated helpful communication and support from their providers. One participant described their provider as “caring” and able to “listen” (P28). Another said they “feel very comfortable in reaching out if there’s any issue … and have open communication,” which they described as “the most important” (P16). Finally, one participant explained how having a long-term relationship with their pharmacist allowed them to get “comfortable saying, ‘Okay, I have this issue’” (P24).

The responsiveness of clinic staff was critical to most participants’ confidence in managing side effects and asking providers for assistance. One participant said, “The doctor’s office is great … They call back” (P2); another described “immediately talking to my doctors when I see something” (P26); one discussed being able to “talk to my doctors about anything” (P35); and one reported feeling “completely fine … telling my doctor about any side effects … and once I figured out what [it was], they figured out a way to deal with it” (P17).

Participants also mentioned “trust” (P30) in their providers as a prominent determinant of adherence. One participant affirmed, “I trusted them well enough to go ahead and try the medication and see if it worked. I would have gone somewhere else to have treatment if I didn’t feel good about the doctor” (P36). Clinicians are critical points of contact for cancer patients, either facilitating or impeding adherence to OAAs. Noting the differences in trustworthiness and communication among providers, one participant conveyed:

> “I don’t know how someone would navigate it without a pharmacist. I saw them alternately: one month, the oncologist, [and] one month, the pharmacist. When I saw the pharmacist, [the pharmacist] would look at my bloodwork in more detail and tell me what things meant. It gave me some peace and some understanding. I feel free to ask questions about the side effects. She [helped] me with some practical suggestions about how to deal with some of the side effects. She was compassionate—is compassionate, still, and it means so much. The oncologist is more detached and preoccupied. I guess it’s the nature of their work. I guess in the scheme of things, my diagnosis is not as dire as some, so I just felt—I feel rushed through, and I don’t feel real compassion, particularly from the doctor. [Nevertheless], the pharmacist has been amazing” (P22).

Overall, participants indicated that compassion and attention to patients’ illnesses encourage trust in providers and facilitate communication about side effects and adherence to OAAs. At the same time, the lack of both contributes to non-adherence.

### Theme 3: “We’re at Their Mercy”: Logistical Factors in Prescription Refills and Shipment

Perhaps another barrier to adherence also comes from the burden of prescription refills and shipment. Although some participants proactively managed their prescription refills and acknowledged that “the onus is on me to make sure that I’m requesting my refills” (P9), many reflected on several challenges with the shipment.

The majority of participants regarded the refill request process as efficient and user-friendly. Participants stated they could easily request refills through digital platforms, which only took a few minutes to complete. One patient summarized the task: “It’s extremely easy because I don’t have to do anything. I just confirm my address with the pharmacist’s office that called, and that’s it” (P1). Another participant shared, “I fill it through CVS Specialty. They usually send me a text a week before I need to fill it out or the week of…When I do see the text, I just click it and fill it. It takes like five minutes” (P3).

While participants generally described requesting refills for their prescriptions as straightforward, several noted difficulties in medication shipment and delivery. These patients expressed frustration with the unpredictability and inconvenience of delivery timelines, particularly the requirement to sign for their medications at home. One patient noted:

> “Well, that’s been the worst. That’s been the most difficult [part of getting] that done. Then, once it is approved, the pharmacy calls [to say] that they got a delivery. Then, I have to make sure that I don’t have an appointment or anywhere to go. Because you have to be home to sign for it” (P31).

Issues also arose when packages were lost, couriers were unable to access neighborhoods, or packages were sent to the wrong address. One patient summarized this burden by saying, “We are at their mercy as far as postal service goes. I think that’s the only concern” (P30).

## Social and economic factors

### Theme 4: “Who can afford that?”: Distress Related to High Costs and Insurance Coverage Challenges

Medication costs and insufficient insurance coverage present further challenges to adherence to OAAs [18]. Many participants emphasized feeling distress due to the high costs of OAA medications. One participant said, “If the [medications] were less expensive … it wouldn’t weigh as heavily on me. I think [that] is the big thing—the cost of the medication” (P10). This participant went on to describe how the “coverage of various … conditions within a health plan … and how much [participants are expected] to cover prior to [the insurance] even kicking in … weighs heavily [on participants]” (P10). The financial strain of OAA caused participants to feel significant distress particularly when they encountered catastrophic out-of-pocket expenses. One participant explained, “I got a notification from my insurance company early on that my medication would not be covered …That first year, the total expense [was] $1,300 a month. That was what it cost me for a 30-day supply: $1,300. I had to pay that out of pocket … because the insurance company wasn’t going to cover it … That’s $1,300 a month times 12 months … Here’s $16,000 in medication. Who can afford that?” (P9).

Sometimes, increasing the medication per shipment mitigates the prescription cost burden slightly.

> “I was getting it every month, and then, to save a little money, he upped it to three months. I get a three-month supply at one time … it saves a little bit. at $100 for a 30-day supply. You know how tight money is” (P6).

Although participants did not report deliberately skipping or reducing doses to save on medication costs, one participant described stretching doses due to an insurance delivery delay.

> “I have, if I had an insurance delivery problem or just a delivery problem…They weren’t able to get it to me in time, so instead of taking two each time, I took one and stretched it out for two days instead of it being one day” (P18).

This highlights how financial and logistical barriers can indirectly contribute to lapses in adherence, even when patients are motivated to stay on treatment.

Patient assistance programs (PAPs) also help alleviate financial toxicities and associated negative impacts. Some of those with financial needs expressed appreciation for PAPs that wholly or partially covered the costs of their drugs, allowing them to maintain adherence to OAAs. One participant said:

> “At $13,000 a month … a cancer foundation pays for my cancer medications. …The only thing I pay for is I pay for my bloodwork, and when I go to see the doctor. Well, it was $25, but now the copay is $15. Then, [for] any other medication that I need, I have copays too, but not the [OAA], thank God…Well, I think it’s because Blue Cross does pay something toward that … No one that I know of could afford $13,000 a month” (P20).

Insurance coverage was not an issue for all participants. One said, “I don’t pay anything. My insurance covers all of it” (P17). Even so, participants generally acknowledged coverage challenges and limited health plans for OAA refills while appreciating comprehensive OAA coverage and patient assistance programs.

### Theme 5: “My wife…knows sometimes I forget”: Family and Social Support in OAAs Adherence

Participants emphasized that family and social support could serve both as barriers and facilitators in maintaining adherence to OAA. They especially expressed how these supports inspire motivation to adhere to treatment. For these participants, motivation was frequently rooted in pursuing a sense of purpose and self-worth, maintaining good health for loved ones, taking care of young or aging family members, and responding positively to consistent support from others.

For most, the desire to remain healthy and continue taking medications was grounded in a deep commitment to loved ones. One patient expressed this powerful motivation: “I get to watch [my child] graduate this year. I made it this far … Living for my kids, that would be the biggest benefit of taking medication” (P12). Another participant articulated a similarly strong sense of duty to be there for their family, underscoring the importance of medication in sustaining their health: “Well, my main reason is I want to be around a long time for my family, and I want to live, and I want to do everything possible that I can, and if taking this medication is [going to] make that happen, that’s what I want to do. I’ve got too much to live for to be [giving] up” (P29).

Family support also meant a regular source of reminder for some participants to adhere to their OAAs. “My kid reminding me. My [wife] reminding me … They’ll just like randomly go, ‘Hey, did you take your medicine?’” (P4).

However, while family support bolstered adherence for several participants, others highlighted how family dynamics could sometimes complicate medication routines. Participants with young children, for example, found it challenging to time their doses precisely due to the demands of family life. “I have a [young child] …[their] routine [bedtime schedule, etc.] made it difficult at times to be consistent” (P5).

Evidently, these participants saw adherence to OAA as sustaining their lives and nurturing meaningful relationships with loved ones.

#### Reciprocal determinism

In sum, the study revealed the different interrelated factors, from personal to healthcare system and social support, that collectively influence OAA adherence. The following excerpt captures and summarizes the above main identified themes and their dynamic interactions.

> “We have been given the time on Earth to live, and you want to live it as comfortably and as normal as you have done in the past. We trust our medical staff to come up with the best solution [for us] at the [right] time. In looking at that and saying, ‘Why do you want to keep going?’ You have family, friends, and purpose in life. Those are things that certainly help motivate [me] to take the medication. I haven’t had any adverse effects that [make me] say, ‘Hey, this is detrimental to me. I can’t take it anymore. It’s killing my body.’ We do know that those things in your body are abnormal, but there were already abnormalities in the body …The meds are trying to combat that” (P10).

## Discussion

Our study went beyond evaluating the known barriers of OAA medication adherence by also characterizing mediators such as self-efficacy and provider communication using Bandura’s Social Cognitive Theory. Prior studies focus on proximal adherence barriers (e.g., side effects, forgetfulness, cost) with little attention given to underlying influences. Our investigation also extends existing knowledge about OAA adherence by concentrating on environmental determinants, especially care team and system-related factors. Our results demonstrate how personal factors and socio-environmental elements influence medication adherence.

We found that patients with high levels of self-efficacy also demonstrated self-reported knowledge and skills in managing their OAAs. The level of self-efficacy determines adherence by directly affecting patients’ ability to manage their health. According to SCT, individuals with high self-efficacy tend to perform behaviors that support long-term health goals. Knowledge enables patients to control their behavior while converting concerns into preventive action. Prior evidence has demonstrated that higher self-efficacy and better disease and treatment knowledge lead to improved medication adherence [19–23]. As such, OAA adherence-centered interventions need to focus on assessing and tracking patient self-efficacy and tailoring approaches according to patient knowledge and understanding.

Consistent with prior studies, we found that interpersonal relationships profoundly influence adherence to medication. Participants reported that caregiver support and responsive provider communication were essential for adherence. Family was the main source of motivation and support for adherence to OAAs for most participants. Razavi and colleagues demonstrated the effective role of social support in increasing treatment adherence in cancer patients [24]. Patients also valued prompt provider responses to side effect concerns and highlighted a need for clear guidance on dose management and drug interactions. Several patients experienced confusion or inadequate support from clinicians, which negatively impacted their trust and symptom management. Patients described being moved across multiple specialists or perceiving differences in care quality between oncologists and pharmacists, underscoring the risks of fragmented care. Together, such experiences illustrate how siloed communication across specialties can leave patients unprepared for the trade-offs imposed by OAA treatment. These gaps highlight the need for more integrated, team-based models of care, such as onco-primary care, that can bridge oncology, primary care, and other specialties to provide holistic guidance across the treatment continuum and lay the foundation for stronger therapeutic relationships [25]. Prior work has also shown that positive provider-patient communication fosters a strong therapeutic alliance and enhances adherence [26–28]. Positive communication builds trust and helps patients feel heard and supported, which is crucial for sustained behavioral engagement. Ultimately, medication adherence is fundamentally rooted in healthy, supportive, communicative, and trusted relationships in patient-related, social/economic, and health care team/system-related domains.

We also found that key environmental barriers, such as cost and shipping coordination, represent major gaps in care. Participants reported frustrations with mail-order pharmacies, including delivery errors and logistical challenges. These issues added stress and reduced patients’ confidence in the healthcare system. Additionally, the high cost of OAAs remains a significant challenge [29–32]. Prior research indicates that cost-related barriers and refill delays adversely impact adherence [29, 33]. Our findings add that prescription shipping logistics are also major underrecognized barriers to sustained OAA use.

In order to address the multiple challenges to OAA adherence identified in this study, an integrated, theory-informed patient support intervention is needed. A multidisciplinary approach involving pharmacists, who are often underutilized in OAA care, can address key barriers such as side effect management and medication coordination while also supporting patient self-efficacy for OAA management. Systematic reviews support the use of personalized interventions delivered by pharmacists as the most effective method to enhance medication adherence [34–39]. Our team is actively developing and testing such an intervention (NCT06989489), grounded in Social Cognitive Theory (SCT) and guided by the Intervention Mapping (IM) framework, and tailored to address individual, interpersonal, and institutional factors [40].

The study provides valuable knowledge about OAA adherence but includes limitations. Self-selection may introduce bias, as individuals willing to discuss their adherence may not fully represent the broader population. Reliance on retrospective reports could introduce potential recall bias. Although our study was reflective of patients receiving care in community facilities of different sizes as well as academic med centers, the shared university affiliation of all sites may somewhat limit generalizability.

Our findings indicated that OAA adherence is shaped by complex interactions across individual, behavioral, and environmental domains, with interpersonal relationships emerging as a particularly powerful influence. Knowledge and self-efficacy serve as essential components, which—together with behavioral skills and social and environmental support—influence adherence. The findings underscore the need to design theory-guided interventions that address multilevel barriers to enhance adherence and, ultimately, improve health outcomes.

## Data Availability

All data produced in the present study are available upon reasonable request to the authors.

