## Supplemental Table 1 for "Determinants of Oral Chemotherapy Adherence: A Social Cognitive Perspective"

**Table 1.** Participant characteristics, stratified by cancer center type.

|  |  | **Facility Type** | | |
| --- | --- | --- | --- | --- |
| **Characteristics** | **Total (N=36)** | **Academic (n=12)** | **Urban (n=12)** | **Rural (n=12)** |
| **Age (years)** |  |  |  |  |
| Median | 62.5 | 57.5 | 61.0 | 69.5 |
| Range | 26-85 | 26-82 | 39-85 | 51-82 |
| **Gender, n (%)** |  |  |  |  |
| Male | 14 (38.9%) | 6 (50.0%) | 3 (25.0%) | 5 (41.7%) |
| Female | 22 (61.1%) | 6 (50.0%) | 9 (75.0%) | 7 (58.3%) |
| **Race/Ethnicity, n (%)** |  |  |  |  |
| White | 29 (80.6%) | 7 (58.3%) | 10 (83.3%) | 12 (100.0%) |
| Black or African American | 5 (13.9%) | 5 (41.7%) | 0 (0.0%) | 0 (0.0%) |
| Asian or Asian American | 1 (2.8%) | 0 (0.0%) | 1 (8.3%) | 0 (0.0%) |
| Other or prefer not to respond | 1 (2.8%) | 0 (0.0%) | 1 (8.3%) | 0 (0.0%) |
| **Cancer Type, n (%)** |  |  |  |  |
| Heme | 23 (63.9%) | 9 (75.0%) | 5 (41.7%) | 9 (75.0%) |
| Solid | 13 (36.1%) | 3 (25.0%) | 7 (58.3%) | 3 (25.0%) |
| **Interview Format, n (%)** |  |  |  |  |
| Individually | 34 (94.4%) | 12 (100.0%) | 11 (91.7%) | 11 (91.7%) |
| With caretaker present | 2 (5.6%) | 0 (0.0%) | 1 (8.3%) | 1 (8.3%) |

*Note:* Academic facility refers to UNC Medical Center, urban refers to UNC Health Rex, and rural refers to McCreary Cancer Center.
