## Supplemental Table 2 for "Determinants of Oral Chemotherapy Adherence: A Social Cognitive Perspective"

**Table 2.** Mapping of WHO Dimensions, SCT Constructs, and Illustrative Quotes

| WHO Dimension | SCT Construct(s) | Illustrative Quotes |
| --- | --- | --- |
| Patient-related factors  *Theme 1: “[I feel] very confident…I’m staying positive about it”: Knowledge, Skills, and Self-Efficacy* | Self-efficacy | “Very confident, very. I haven't had anything yet that has come across that I haven't been able to handle. I feel like the medication is really doing its job.” (P2) • “Oh, real confident. Very confident [managing side effects] …Yes. Definitely [feel comfortable asking my doctor for help with side effects] …my doctors I could talk to about anything …” (P35) |
|  | Behavioral capability | “I keep them in a bag I take with me. Yeah…Right [they’re always with me].” (P18) • “I take all my medications right before I go to bed at night, so it's just part of my nightly routine. When I go to brush my teeth and wash my face, I take my medicine.” (P23) |
|  | Expectations | “Medically it's designed to address a cancer, so either person wants to live, or they don't. I happen to want to. That's a motivator. That's the only motivator. Other than that, I don't like taking medicine.” (P1) • “I'm motivated to take my medication because I don't want to die. That's all. I figured, the chemo pills would help me, and that was the motivation of taking it.” (P15) • “Wow. I'm still alive when, if this had been not that long ago, I would have already died. The benefits are endless. I'm alive, and not only alive, but I'm alive and doing well. I'm able to function. I don't know. That's definitely the benefit (P2). |
| Health care team & system-related factors  *Theme 2: “They’ll stop me from speaking”: Communication and Trust with Providers* |  |  |
|  | Reinforcement | “… Accredo is a direct mail, and they're amazing. They call me once a month, to make sure nothing in my health has changed. They ask me if I would like to speak with a pharmacist. Then, when we agree that I don't need to speak with a pharmacist and everything is like it was, that I'm not on any new medications, they just automatically send it to me, and it gets delivered to my house by Federal Express. (P2) • “I think working with a patient to really help them sort through their routine…once you have a routine, the stress is improved.” (P22) |
| *Theme 3: “We’re at Their Mercy”: Logistical Factors in Prescription Refills and Shipment* |  |  |
|  | Reciprocal determinism | “I think having the bigger supply helps me, because there would be times, I'm sure that I was like, oh, my gosh, I meant to go pick that up, and the pharmacy's closed, and now I have to wait until tomorrow. I think that option for that happening less, every 90 days, instead of every 30 days, definitely helps me.” (P23) |
| Social & economic factors  *Theme 4: “Who can afford that?”: Distress Related to High Costs and Insurance Coverage Challenges* | Reciprocal determinism | “We had to cut back on groceries some months to afford my medicine.” (P20) • “The bills come at the same time as everything else—utilities, rent—it all piles up.” (P16) |
| *Theme 5: “My wife…knows sometimes I forget”: Family and Social Support in OAA Adherence* | Observational learning | “… I would say I would encourage them mostly to find a support group of people that can easily share information easily. Not waiting for a once-a-month meeting, or a once-a-week meeting. I'm talking about an internet-type share, because that's what I have found to be the very most helpful thing, of my entire experience, is having those people that I can quickly put on a message board, and people from all over the world can give their input and advice.”(P2) • “… some recommendations, just glowing recommendations from other patients and their families.” (P36) |
|  | Reciprocal determinism | “My husband is [helping me with medication]. He helps me with it. If I'm not gotten up or somethin' and gotten it out already, when he gets up, he'll see that it's not out, and he'll go ahead and get it out.” (P29) • “… because my wife is working. We are from—originally from Morocco. We don’t have no family over here. It’s just like some friends, but everybody has his own things… it’s … a private company [that employees the person that comes to help him each day], was hired by the insurers. I think. … they help with everything. They make sure for your safety and to go to restrooms and come back and those things. The food—if you need any food, and any other things. Daily life…It is nice.” (P21) |
